# Histomorphometric features of placentae from women having malaria and HIV coinfection with preterm births

**DOI:** 10.1101/2023.10.30.23297751

**Authors:** Khalil Y. Adam, Obimbo M. Moses, Gitaka Jesse, Walong Edwin, Ogutu Omondi, Stephen.B.O. Ojwang

**Affiliations:** Department of Obstetrics and Gynaecology, University of Nairobi. Kenya; Department of Human Anatomy and Physiology, University of Nairobi. Kenya; College of Health Sciences, Mount Kenya University. Kenya; Department of Human Pathology, University of Nairobi. Kenya; Basic Clinical and Translational Research Laboratory, Nairobi. Kenya

**Keywords:** Preterm Birth, Malaria, HIV Infection, Pregnancy Outcome, Placenta, Maternal Vascular Malperfusion

## Abstract

**Background:** Malaria and HIV are associated with preterm births possibly due to partial maternal vascular malperfusion resulting from altered placental angiogenesis. There is a paucity of data describing structural changes associated with malaria and HIV coinfection in the placentae of preterm births thus limiting the understanding of biological mechanisms by which preterm birth occurs.

**Objectives:** This study aimed to determine the differences in clinical characteristics, placental parenchymal histological, and morphometric features of the terminal villous tree among women with malaria and HIV coinfection having preterm births.

**Methods:** Twenty-five placentae of preterm births with malaria and HIV coinfection (cases) were randomly selected and compared to twenty-five of those without both infections (controls). Light microscopy was used to determine histological features on H&E and MT-stained sections while histomorphometric features of the terminal villous were analyzed using image analysis software. Clinical data regarding maternal age, parity, marital status, level of education, gestational age and placental weight were compared.

**Results:** Placental weight, villous perimeter and area were significantly lower in cases as compared to controls 454g vs. 488g, 119.32µm vs. 130.47µm, and 937.93µm^2^ vs. 1132.88µm^2^ respectively. Increased syncytial knots and accelerated villous maturity were significantly increased in the cases. The relative risk of development of partial maternal vascular malperfusion was 2.1 (CI: 1.26-3.49).

**Conclusion:** These findings suggest that malaria and HIV coinfection leads to partial maternal vascular malperfusion that may lead to chronic hypoxia in the placenta and altered weight, villous perimeter and surface area. This may represent a mechanism by which malaria and HIV infection results in pre-term births.

## Introduction

Preterm birth is defined as birth occurring before the completion of 37 weeks of gestation. According to the World Health Organisation (WHO), preterm birth is a global health challenge and a significant contributor to neonatal mortality and morbidity (1), with South Asia and Sub-Saharan Africa accounting for over 60% of cases. It is estimated that preterm births account for 35% of global neonatal deaths (2). While national estimates for African countries with reliable data are varied, in Kenya, the prevalence of pre-term birth at the national referral hospital has been estimated as 18.3%(3–5) highlighting the need for escalating antenatal care to mitigate the clinical sequelae of pre-term births.

The aetiology of pre-term birth is multifactorial and the mechanisms are still not well understood. Medical conditions in pregnancy, uterine and cervical anomalies and infections in pregnancy are some of the known causes. Infection with Malaria and HIV during pregnancy has been associated with PTB. The incidence of preterm birth is higher in Malaria (44.6%) and HIV infection (33.4%) compared to uninfected controls (6). Importantly, up to 37% of HIV-infected pregnant women in Sub Saharan Africa contract malaria, and the incidence of preterm birth in this coinfection is likely to be higher due to the bidirectional and synergistic relationship between the two infections (7,8).

One possible mechanism of preterm birth in malaria and HIV coinfection is partial maternal vascular malperfusion resulting from the altered angiogenesis in the decidual arterioles of the placenta (9,10). This results in reduced and abnormal flow of maternal blood in the intervillous space and ultimately a state of chronic placental hypoxia. Histologically, partial maternal vascular malperfusion presents identifiable changes categorized as accelerated villous maturity and distal villous hypoplasia (11–13). Partial maternal vascular malperfusion is associated not only with occurrence but also recurrence of preterm births, and its diagnosis has implications on the management on future pregnancies (13).

Despite increased epidemiological and clinical research on the interaction between malaria, HIV and PTB (14) studies investigating the potential biomechanisms underlying these observations are still lacking (15). This study aimed to determine the differences in clinical characteristics, histological and morphometric features of placental parenchyma of the terminal villous tree among preterm births in women with malaria and HIV coinfection and those without.

## Materials and Methods

### Study population

This retrospective cohort study was designed to analyze the differences in the histology and morphometry of preterm placental parenchyma in women with malaria and HIV coinfection compared to those without. Ethical approval for the study was obtained from the Kenyatta National Hospital–University of Nairobi Ethics and Review Committee (KNH/UON ERC), reference number P406/08/2020. Placenta specimens were obtained from the biobank at the Basic Clinical and Translational Research Laboratory, Department of Human Anatomy and Physiology, University of Nairobi. These samples had been collected between January 2018 and December 2019 at the Bungoma County and Referral Hospital. The hospital has an average of 600 deliveries monthly and is located in Western Kenya, an area with a high prevalence of both malaria and HIV (16–18). Placentae specimens included in this study were from women aged between 18 to 40 years who had a singleton pregnancy. Gestational age included was 28 weeks plus 0 days to 36 weeks plus 6 days as calculated from their first day of the last menstrual periods or estimated by ultrasound scan before 22 weeks. Those with hypertension, diabetes, anaemia, malnutrition, chorioamnionitis, preterm premature rupture of membranes, and other infections other than malaria and HIV were excluded. The clinical and sociodemographic data were obtained from clinical data forms that were used to abstract data from patient records. An online random number generator (https://numbergenerator.org/randomnumbergenerator) was used to select specimens from the sampling frame of the biobank. Only placenta blocks obtained from the central part of the normally shaped placentae were analysed in this study.

### Specimen preparation and staining

From 50 placentae blocks (25 in each group), six 5µm serial sections were made using the Leitz Wetzler sledge microtome. These sections were floated in a water bath at 40°C, then fished out on glass slides and dried overnight in a dry heat oven at 40°C. Each section was then deparaffinized and rehydrated in decreasing alcohol concentration and stained with Haematoxylin and Eosin (H&E) and Masson’s Trichome (MT), after which they were dehydrated in increasing concentrations of alcohol and mounted on slides with DPX (Dibutylphthalate Polystyrene Xylene). Two slides were randomly selected for light microscopy (one slide stained by H&E and the other slide stained by MT), giving a total of 100 slides analyzed by light microscopy (50 H&E and 50MT). From each of the H&E-stained slides, three microphotographs were captured from random fields, and a total of 150 microphotographs were analysed morphometrically.

### Microscopy

The Richter Optica XU 1T plan digital microscope (manufacturer, city, country) interphased with Moticam BTU 10 digital camera (manufacturer, city, country) was used for light microscopy. Analysis was done at a total magnification of 100X and 400X. Distal villous hypoplasia and callous vascularity were analyzed at a total magnification of 100X. Diagnosis of chorangiosis was made after demonstrating 0 0r more capillaries per villous in more than 10 villi in 10 fields seen under X10 magnification., while the overall slide diagnosis was made in consensus with the Amsterdam Criteria 2014 (13,19). hypervascularity, normal vascularity, hypervascularity and chorangiosis were defined as one or less, two to six, seven to ten and more than ten capillaries per villous.

Microphotographs were captured at a resolution of 1280×800 pixels at a total magnification of 400X by the Moticam BTU10 camera system, which was calibrated to a known scale (µm) and saved as BMP files. The Fiji® image processing package version 2.1.0. (manufacturer/vendor, city, country) was used to estimate the villous diameter (µm), perimeter (µm), and cross-sectional area (µm^2^) of the terminal villous as follows; the images were first opened in Fiji® version 2.1.0. (20) then converted to an 8-bit image, scaled in µm, segmented by auto-threshold and measured.

### Statistical analysis

Data were analyzed using Statistical Package for Social Scientist (SPSS) (Version 26.0, Chicago, Illinois). Numerical data were analyzed using the Student’s t-test or Mann Whitney U test for normally and non-normally distributed data respectively. Categorical data were analyzed using the X^2^ test or Fisher’s Exact test. Relative Risk was calculated to measure the strength of association between partial maternal vascular malperfusion/villous hypervascularity and malaria and HIV coinfection. For all statistical analyses, a p-value of ≤0.05 was considered significant.

## Results

### Clinical and Sociodemographics

A total of 50 women were included in this study, twenty-five (25) of whom had malaria and HIV coinfection while 25 tested negative for both infections and were in the comparison group. The mean maternal age and parity were significantly lower in women with malaria and HIV coinfection (26 years vs. 29 years and 2 vs. 3, respectively). The level of education was also significantly lower in women with malaria and HIV coinfection compared to those without with most having primary education. There was no difference in the mean gestational age and marital status between the two groups (see table 1).

**Table 1:** Clinical and sociodemographic characteristics of women delivering preterms. HIV-Human immunodeficiency virus; m-mean; SD-standard deviation.

| Clinical and social demographic features | Malaria and HIV coinfection<br>n=25 | Malaria and HIV negative<br>n=25 | p-value |
| --- | --- | --- | --- |
| **Age(years)[m(SD)]** | 26(5.147) | 29(4.482) | 0.029<sup>α</sup> |
| **Age categories(years)** |  |  |  |
| 15-24 [n(%)] | 10(40%) | 4(16%) | 0.059<sup>β</sup> |
| 25-40 [n(%)] | 15(60%) | 21(84%) |  |
| **Parity [m(SD)]** | 2(1.173) | 3(0.997) | 0.012<sup>α</sup> |
| **Parity categories** |  |  |  |
| Primipara [n(%)] | 8(32%) | 0(0%) | 0.004<sup>γ</sup> |
| Multipara [n(%)] | 16(64%) | 23(92%) |  |
| Grand Multipara [n(%)] | 1(4%) | 2(8%) |  |
| **Gestational age at delivery(weeks) [m(SD)]** | 34(1.159) | 35(1.118) | 0.113<sup>α</sup> |
| **Level of education** |  |  |  |
| Primary school | 12(48%) | 2(8%) | 0.007<sup>γ</sup> |
| High School | 9(36%) | 15(60%) |  |
| College | 4(16%) | 8(32%) |  |
| *α- Student t-test* |  |  |  |
| *β- Chi-square test* |  |  |  |
| *γ-Fisher Exact test* |  |  |  |

### Histological features of preterm placentae

The histological features of placental parenchyma were assessed to compare the impact of malaria and HIV coinfection on preterm placentae. There were significantly higher rates of accelerated villous maturity (Figure 1A), increased villous vascularity (Figure 1B), syncytial knots changes, and villous vascularity in the placentae with malaria and HIV coinfection compared to those without. Fibrin deposition was, however, significantly higher in malaria and HIV coinfection group (21/25[84%]) compared to those without (5/25[20%]) (Figure 1E). Distal villous hypoplasia was also more prevalent in malaria and HIV coinfection group although the difference was not statistically significant. Photomicrographs showing the distal villous hypoplasia and normal villous density are shown in Figures 1C and 1D. Villous necrosis and villous stromal fibrosis were also observed in 4/25(16%) placentae in the group without malaria and HIV coinfection, this feature was not seen in malaria and HIV co-infection group. (Figure 1F). The overall impression of partial maternal vascular malperfusion was significantly higher in malaria and HIV coinfection group compared to those without (Table 2).

**Figure 1.**
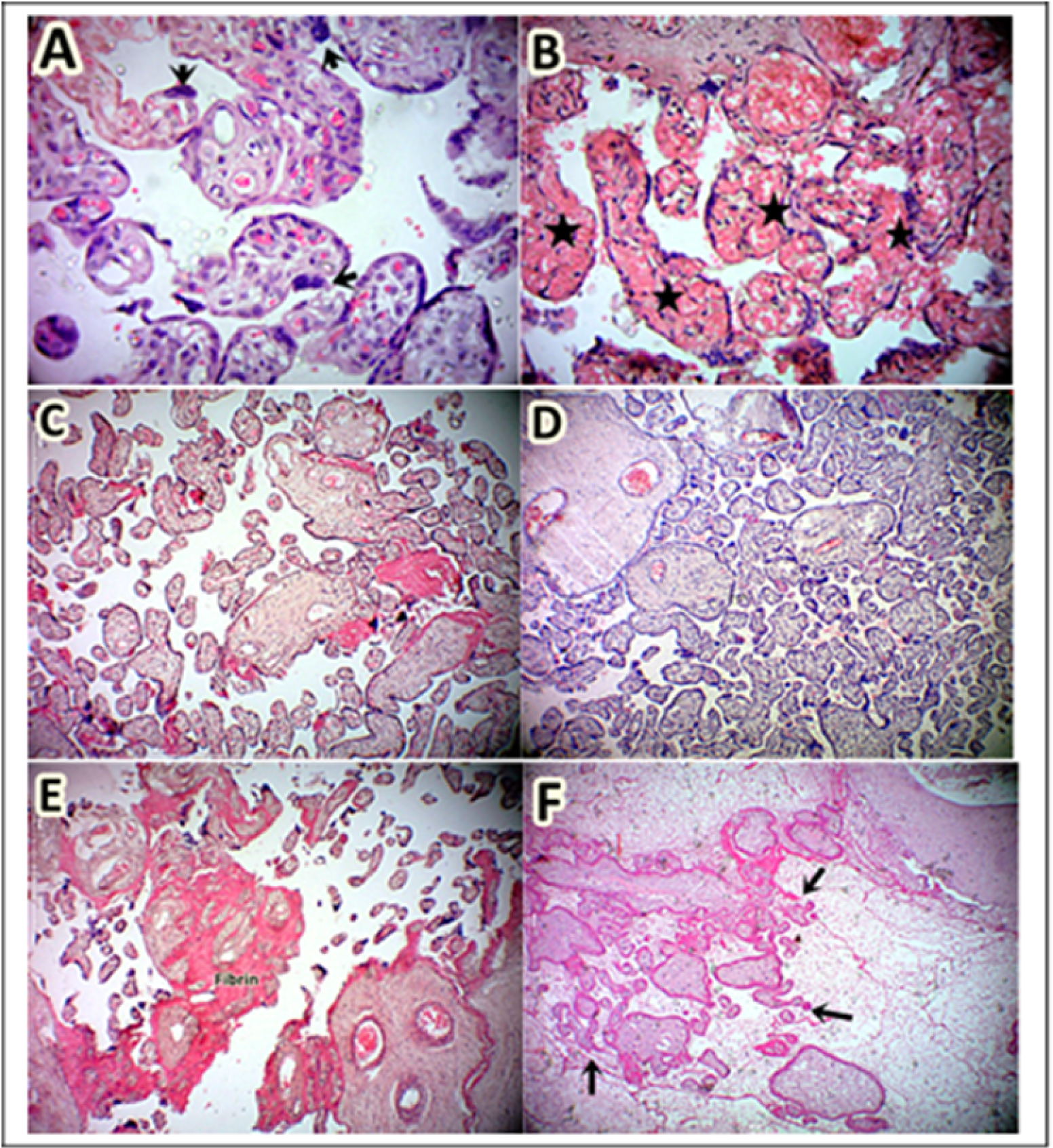
Histological features of placental parenchyma showing features of maternal vascular malperfusion. **A.** Increased vesciculosyncitial membrane and reduced terminal villous dimension suggestive of accelerated villous maturity, **B.** Increased villous hypervascularity, **C.** Scarcity of terminal villous compared to the intervillous space indicative of distal villous hypoplasia, **D.** Normal terminal villous concentration, **E.** Increased fibrin deposition, and F. Villous necrosis.

**Table 2:**
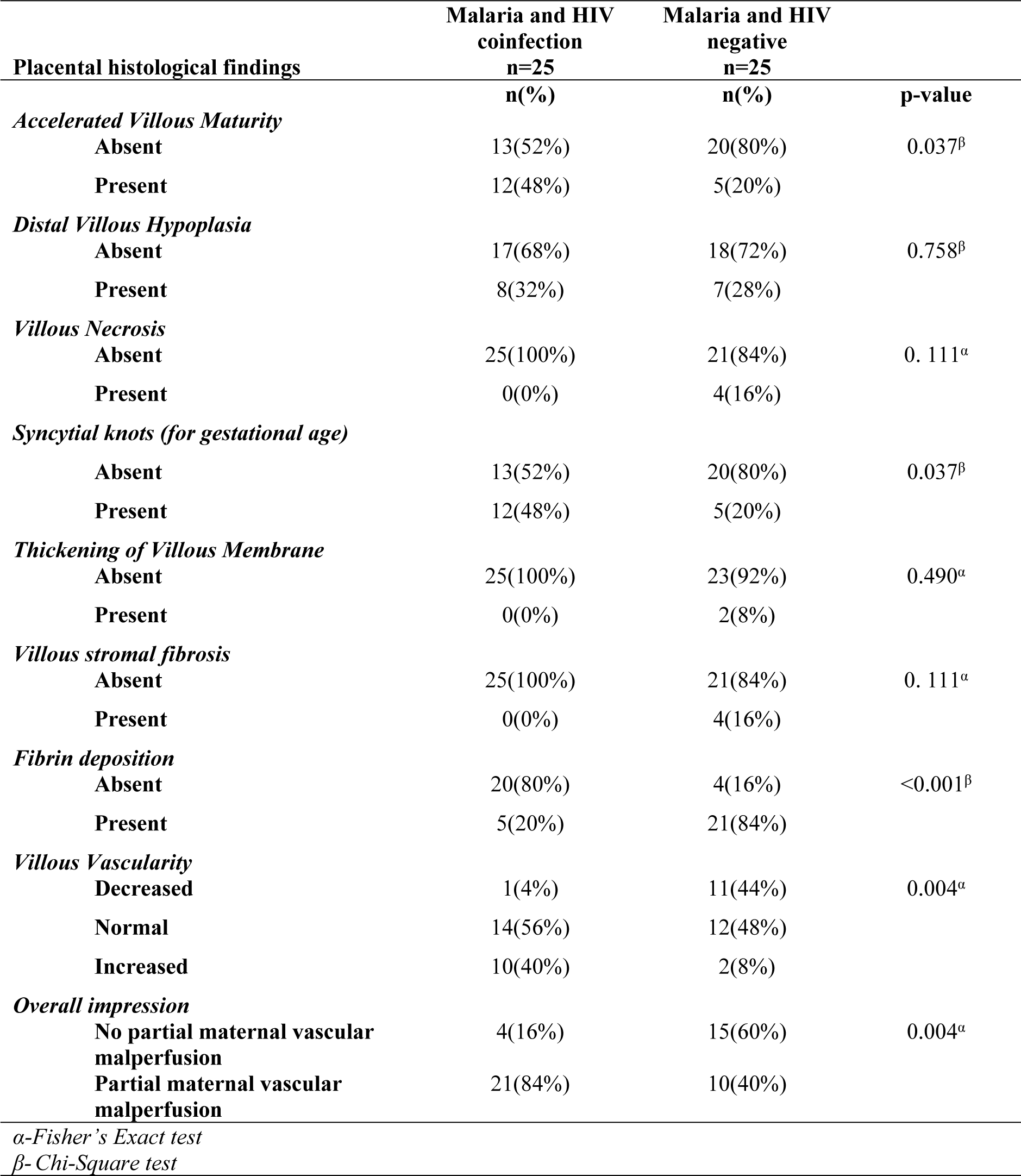
Comparison of the histological features of placental parenchyma among preterm placentae. HIV-Human immunodeficiency virus.

### Histomorphometric features of preterm placentae

The preterm placentae with malaria and HIV coinfection were compared to those without the coinfection to determine the differences in histomorphometric features. The mean placental weight, the perimeter and cross-sectional area of the terminal villi were significantly lower in malaria and HIV group compared to the group without the coinfection (454.4±32.02 g vs 488±36.74 g), (119.32µm vs. 130.47µm) and (937.93µm^2^ vs. 1132.88µm^2^), respectively. The diameter was lower, while the average capillary number per terminal villous was higher in malaria and HIV coinfection group compared to those without, although the difference was not statistically significant (Table 3).

**Table 3:** Morphometrical parameters of the terminal villi among preterm placentae. HIV-Human immunodeficiency virus; m-mean; SD-standard deviation.

| histomorphometric features | Malaria and HIV<br>coinfection<br>n=25 | Malaria and HIV<br>negative<br>n=25 | p-value |
| --- | --- | --- | --- |
|  | *m(SD)* | *m(SD)* |  |
| Placental weight(g) | 454(32) | 488(36.7) | 0.001<sup>a</sup> |
| Diameter ($\mu\text{m}$) | 41.24(4.406) | 43.37(4.611) | 0.102<sup>a</sup> |
| Perimeter ($\mu\text{m}$) | 119.32(9.2) | 130.47(12.47) | 0.001<sup>a</sup> |
| Cross sectional area of villous ($\mu\text{m}^2$) | 937.93(148.6) | 1132.88(235.85) | 0.001<sup>a</sup> |
| No of capillaries / villous | 5.28(3.93) | 3.24(2.48) | 0.099<sup>b</sup> |
| *α-Student t-test* |  |  |  |
| *β-Mann Whitney U test* |  |  |  |

### Risk of maternal vascular malperfusion

To determine the risk of maternal vascular malperfusion in the preterm placentae of malaria and HIV coinfection group, a 2 by 2 table with Relative Risk was used and the relative risk of having partial maternal vascular malperfusion was significantly higher in preterm births with malaria and HIV coinfection than those without; RR 2.10 CI (1.26-3.49) (Table 3). For villous hypervascularity, the RR was 4 CI (0.94-17.00) (Table 4)

**Table 4:**
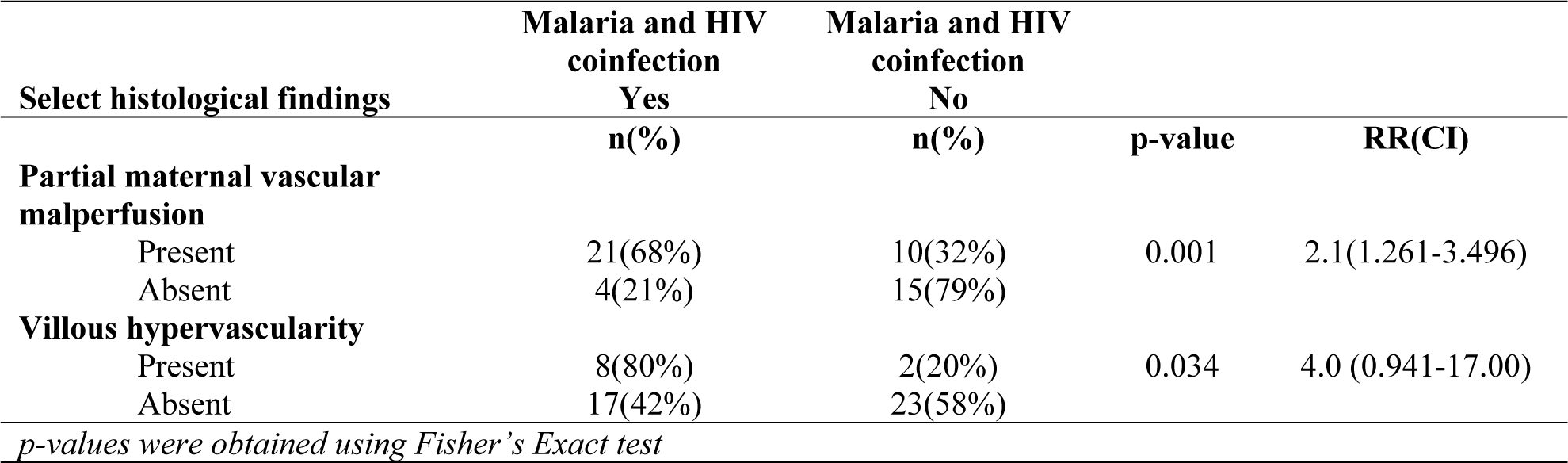
Relative Risk of partial maternal vascular malperfusion and villous hypervascularity among preterm placentae. HIV-Human immunodeficiency virus; RR-relative risk; CI-confidence interval.

## Discussion

In this study, we investigated the impact of malaria and HIV coinfection on the structire of preterm placentae. We compared the clinical characteristics and microscopic structure of the placental parenchyma between preterms born to mothers having malaria infection and living with HIV to those born to mothers without the coinfection. We observed, that women with malaria and HIV coinfection were younger, of lower parity and level of education and that their placentae had increased risk of having maternal vascular malperfusion.

Malaria and HIV tend to be more prevalent in the young age groups and women with low parity and level of education. Women of young age are particularly vulnerable to HIV because of the risky behaviour including unprotected sex and drug use (21). It is also known that naturally acquired immunity to malaria increases with increasing parity (22,23). Education is important in understanding the disease in terms of transmission, treatment and prevention (24). Our study supports these findings and therefore we underscore the importance of increasing public health intervention aimed at increasing the level of education of the young age.

A number of studies have investigated placental structures separately in HIV and malaria. Features of partial maternal vascular malperfusion such as increased syncytial knotting - a component of accelerated villous maturity - have been observed in placentae of women with HIV and malaria infections (11,12,25). Given that sub-Saharan Africa has a high burden of malaria and HIV infection, it is plausible that these two infections may have a synergistic effect on placental histology. (7,8). Our study found that increased syncitial knotting and accelerated villous maturity is significantly higher in malatria and HIV coinfection and therefore, supports the hypothesis that maternal vascular malperfusion could be an important biological mechanism that xcmodulates placental changes thereby contributing to preterm births in malaria and HIV coinfection. Indeed, we found that the relative risk of having maternal vascular malperfusion was 2.1, suggesting that having malaria and HIV are significant risk factors for development of MVP during gestation.

Studies have shown that partial maternal malperfusion results from inflammation and altered angiogenesis from the two infections and this ultimately leads to a chronic hypoxic state that results in adverse pregnancy outcomes including preterm birth (10,26–28). Malaria and HIV infections potentially promote the antiangiogenic state by reducing angiopoietin 1 and placental growth factor production and increasing soluble endoglin levels (29–31). Intervillous inflammation and decidual vasculopathy in these infections may cause occlusion of decidual maternal vessels that leads to ischemia and hypoperfusion. While we did not study these biomarkers, our study highlights the need for more studies to investigate the longitudinal changes in the levels of these biomarkers throughout the gestation period. Such studies would be useful in creating predictive models of adverse pregnancy outcomes including preterm births. Furthermore, the discovery of potential biomarkers would facilitate the development of rapid diagnostic kits for early detection of high risk pregnancies. Moreover, such studies may lead to the discovery or repurposing of drugs that can halt or reverse these detrimental changes and reduce the burden of adverse outcomes in pregnancies affected by malaria, HIV and other medical conditions.

An important clinical implication of the presence of maternal vascular malperfusion is its association with recurrence of adverse pregnancy outcomes in subsequent pregnancies. These adverse outcomes are collectively referred to as ischaemic placental diseases and include preterm births, preeclampsia and fetal growth restriction. Ou finding of maternal vascular malperfusion in pregnancies affected with malaria and HIV justifies the employment of interventions that are known to prevent or reduce the incidences of adverse maternal outcomes, for example early antenatal care, frequent visits, and fetal surveillance, aspirin therapy, early delivery after antenatal corticosteroids, and optimization of maternal cardiovascular function by controlling blood pressure and diabetes (13,32). In the present study, all participants were treated appropriately for both conditions according to national clinical guidelines. Neither the typical histological findings seen in malaria, namely hemozoin deposition and intervillositis, nor the villitis in HIV infection was observed. This finding suggests that treatment had clinical and histological benefits as has been similarly reported in past investigations (33,34) and underpins the integral role of optimal treatment and adherence counselling for malaria and HIV during antenatal care for co-infected patients.

To the best of our knowledge, this is the first study in Kenya to study placenta structure in preterm birth among women with malaria and HIV coinfection. We used the terminologies laid out by the Amsterdam Consensus Group for placental lesions hence this study can be compared to other using the same terminologies. We also acknowledge some limitations to our study. Although our study was powered to compare the differences in the two study groups, our analysis of clinical characteristics was limited to maternal age, parity, level of education as these were the only available data that could be obtained for the biobank specimens that we used. The important history of previous adverse pregnancy outcomes including preterm was not obtained as well as information regarding the duration of the disease, level of parasitemia, viral load, CD4 count. A prospective study design would ensure the collecting of all the relevant clinical data needed. Since this was a retrospective study, we did not have access to data on disease duration, treatment regimen and history of preterm births in the clinical form. For the women living with HIV, we did not have data on CD4+ T cell count and viral load at time of specimen collection, which would have been important in determining the impact of immune status and viral dynamics on placental histology. This study was designed to explore the placental histological feature post-delivery and therefore we cannot definitively prove causality. However, a prospective design may pose ethical and feasibility challenges.

In conclusion, this study demonstrates that malaria and HIV co-infection increases the risk of placental histologic changes indicative of malperfusion and chronic hypoxia. These histologic changes may represent an important biological mechanism that underlie the higher rate of preterm births during this coinfection. Placental histology may therefore be essential in the evaluation of the etiology of preterm births and in determining the risk of recurrence. These findings are important in guiding the management of future pregnancies to avoid adverse outcomes in women with HIV and malaria.

## Data Availability

All data produced in the present study are available upon reasonable request to the authors

## Acknowledgement

The authors are grateful to the Staff at the Basic Clinical and Translation Research Laboratory, University of Nairobi for providing and allowing analysis of the bio-specimens. The authors also thank the Staff of the Department of Obstetrics and Gynecology, Department of Human Anatomy and Physiology and the Department of Human Pathology for their input at the various stages of implementing this research.

The Authors also wish to acknowledge the training and support from the University of Nairobi’s Building Capacity for Writing Scientific Manuscripts (UANDISHI) Program at the Faculty of Health Sciences. This work was funded in part by the ADVANCE program at IAVI with support from University of California, San Francisco’s International Traineeships in AIDS Prevention Studies (ITAPS, U.S. NIMH, R25MH123256) and made possible by the support of many donors, including United States Agency for International Development (USAID). The full list of IAVI donors is available at http://www.iavi.org. The contents of this manuscript are the responsibility of the authors and do not necessarily reflect the views of USAID or the US Government.

## Author Contributions

Khalil YA designed and carried out the study, performed experiments and wrote the article. Obimbo MM participated in the study design, supervised the study and coordinated sample preparation. Walong E performed and reviewed histological diagnoses. Gitaka J, Ogutu O and Ojwang SBO jointly supervised the study. All authors participated in interpretation of data and critical review of the manuscript.

## References

1. Howson CP, Kinney M V, Lawn JE. March of dimes, PMNCH, save the children, WHO. Born too soon: the global action report on preterm birth. Geneva: World Health Organization. 2012.

2. Blencowe H, Cousens S, Oestergaard MZ, Chou D, Moller AB, Narwal R, et al. National, regional, and worldwide estimates of preterm birth rates in the year 2010 with time trends since 1990 for selected countries: A systematic analysis and implications. Lancet [Internet]. 2012;379(9832):2162–72. Available from: 10.1016/S0140-6736(12)60820-4

3. WHO. Preterm Birth [Internet]. 2018. Available from: https://www.who.int/news-room/fact-sheets/detail/preterm-birth

4. Wagura PM. Prevalence and Factors Associated With Preterm Birth At Kenyatta National Hospital. BMC Pregnancy Childbirth. 2018;18(107):2–9.

5. Otieno P, Waiswa P, Butrick E, Namazzi G, Achola K, Santos N, et al. Strengthening intrapartum and immediate newborn care to reduce morbidity and mortality of preterm infants born in health facilities in Migori County, Kenya and Busoga Region, Uganda: a study protocol for a randomized controlled trial. Trials. 2018;19(313):1–12.

6. Juma EO, Keraka M, Wanyoro A. Clinical Phenotypes Associated With Preterm Births at Jaramogi Oginga Odinga Teaching and Referral Hospital in Kisumu County, Kenya. Int J Curr Asp. 2019;3(III):175–86.

7. Kwenti TE. Malaria and HIV coinfection in sub-Saharan Africa: prevalence, impact, and treatment strategies. Res Rep Trop Med. 2018;9:123–36.

8. Kirinyet JK. An Assessment of Malaria Parasite Density among HIV / AIDS-Subjects at Different Levels of CD4 T-Cells Prior to Antimalarial Therapy at Chulaimbo Sub-County Hospital, Western Kenya. J Trop Med. 2019;2019:1– 7.

9. Ataíde R, Murillo O, Dombrowski JG, Souza RM. Malaria in Pregnancy Interacts with and Alters the Angiogenic Profiles of the Placenta. PLoS Negl Trop Dis. 2015;1–15.

10. Conroy AL, Mcdonald CR, Gamble JL, Olwoch P, Natureeba P, Cohan D, et al. Altered Angiogenesis as a common mechanism underlying preterm birth, small for gestational age, and stillbirth in women living with HIV. Am J Obstet Gynecol [Internet]. 2017;217(6):684.e1-684.e17. Available from: 10.1016/j.ajog.2017.10.003

11. Obimbo MM, Zhou Y, Mcmaster MT, Cohen CR, Qureshi Z, Ong J, et al. Placental Structure in Preterm Birth Among HIV-Positive Versus HIV-Negative Women in Kenya. J Acquir Immune Defic Syndr. 2019;80(1):94–102.

12. Ahenkorah J, Tetteh-quarcoo PB, Nuamah MA, Bentum BK, Nuamah HG, Hottor B, et al. The Impact of Plasmodium Infection on Placental Histomorphology: A Stereological Preliminary Study. Infect Dis Obstet Gynecol. 2019;2019:1–8.

13. Redline RW. Classification of placental lesions. Am J Obstet Gynecol. 2015;21–8.

14. Visser L, Boer MA De, Groot CJM De. Analysis of Publication Interest on Preterm Birth Over Two Decades. Matern Child Health J [Internet]. 2019;23(10):1392–9. Available from: 10.1007/s10995-019-02772-x

15. Hochman S, Kim K. The Impact of HIV and Malaria Coinfection: What Is Known and Suggested Venues for Further Study. Interdiscip Perspect Infect Dis. 2009;2009:1–8.

16. JamiiForums [Internet]. 2019. Available from: https://r.search.yahoo.com/_ylt=A0geKYkdnGJiTOcAcRtXNyoA;_ylu=Y29sbwNiZjEEcG9zAzEEdnRpZANMT0NVSTAxOF8xBHNlYwNzYw--/RV=2/RE=1650658462/RO=10/RU=https%3A%2F%2Fwww.jamiiforums.com%2Fthreads%2Fkenya-begins-construction-of-a-maternity-hospital-in-bungoma.16114

17. Zhou G, Zhong D, Lee M-C, Wang X, Atieli HE, Githure JI, et al. Multi-Indicator and Multistep Assessment of Malaria Transmission Risks in Western Kenya. Am J Trop Med Hyg [Internet]. 2021 Feb 8;104(4):1359–70. Available from: https://pubmed.ncbi.nlm.nih.gov/33556042

18. Borgdorff MW, Kwaro D, Obor D, Otieno G, Kamire V, Odongo F, et al. HIV incidence in western Kenya during scale-up of antiretroviral therapy and voluntary medical male circumcision: a population-based cohort analysis. lancet HIV. 2018 May;5(5):e241–9.

19. Srinivasan AP, Omprakash BOP, Lavanya K, Subbulakshmi Murugesan P, Kandaswamy S. A prospective study of villous capillary lesions in complicated pregnancies. J Pregnancy. 2014;2014.

20. Schindelin J, Arganda-Carrera I, Frise E, Verena K, Mark L, Tobias P, et al. Fiji - an Open platform for biological image analysis. Nat Methods. 2009;9(7).

21. Simon Kiprono Ruttoh, Milward Tobias, Benard Kipngeno Ruttoh. The Status of Human Immuno-deficiency Virus (HIV) Infection among Youth Aged 15-24 Years in Malawi and Kenya. J Environ Sci Eng B. 2017;6(7):380–6.

22. Nnaji GA, Okafor CI, Ikechebelu JI. An evaluation of the effect of parity and age on malaria parasitaemia in pregnancy. J Obstet Gynaecol (Lahore) [Internet]. 2006 Jan 1;26(8):755–8. Available from: 10.1080/01443610600956089

23. Duffy PE. Plasmodium in the placenta: parasites, parity, protection, prevention and possibly preeclampsia. Parasitology. 2007;134(Pt 13):1877–81.

24. Tuntufye SM. Education level and human immunodeficiency virus (HIV)/acquired immune deficiency syndrome (AIDS) knowledge in Kenya. J AIDS HIV Res. 2014;6(2):28–32.

25. Chaikitgosiyakul S, Rijken MJ, Muehlenbachs A, Lee SJ, Chaisri U, Viriyavejakul P, et al. A morphometric and histological study of placental malaria shows significant changes to villous architecture in both Plasmodium falciparum and Plasmodium vivax infection. Malar J. 2014;13(4):1–13.

26. Weckman AM, Ngai M, Wright J, McDonald CR, Kain KC. The impact of infection in pregnancy on placental vascular development and adverse birth outcomes. Front Microbiol. 2019;10(AUG):1–11.

27. Eloundou SN, Lee JY, Wu D, Lei J, Feller MC, Ozen M, et al. Placental malperfusion in response to intrauterine inflammation and its connection to fetal sequelae. PLoS One. 2019;14(4):1–15.

28. Visser L, van Buggenum H, van der Voorn JP, Heestermans LAPH, Hollander KWP, Wouters MGAJ, et al. Maternal vascular malperfusion in spontaneous preterm birth placentas related to clinical outcome of subsequent pregnancy. J Matern Neonatal Med [Internet]. 2019;0(0):1–6. Available from: 10.1080/14767058.2019.1670811

29. Graham SM, Rajwans N, Tapia KA, Jaoko W, Estambale BBA, McClelland RS, et al. A prospective study of endothelial activation biomarkers, including plasma angiopoietin-1 and angiopoietin-2, in Kenyan women initiating antiretroviral therapy. BMC Infect Dis [Internet]. 2013;13(1):1. Available from: BMC Infectious Diseases

30. Motomura K, Romero R, Garcia-Flores V, Leng Y, Xu Y, Galaz J, et al. The alarmin interleukin-1a causes preterm birth through the NLRP3 inflammasome. Mol Hum Reprod. 2020;26(9):712–26.

31. De Jong GM, Slager JJ, Verbon A, Van Hellemond JJ, Van Genderen PJJ. Systematic review of the role of angiopoietin-1 and angiopoietin-2 in Plasmodium species infections: biomarkers or therapeutic targets? Malar J. 2016;15(1):1–12.

32. Ernst LM. Maternal vascular malperfusion of the placental bed. Apmis. 2018;126(7):551–60.

33. Bruce-Brand C, Schubert PT, Wright CA. HIV, placental pathology and birth outcomes - a brief overview. J Infect Dis [Internet]. 2021; Available from: http://europepmc.org/abstract/MED/33987644

34. Muehlenbachs A, Nabasumba C, McGready R, Turyakira E, Tumwebaze B, Dhorda M, et al. Artemether-lumefantrine to treat malaria in pregnancy is associated with reduced placental haemozoin deposition compared to quinine in a randomized controlled trial. Malar J. 2012;11:1–9.

